# Population Structure, Novel Sequence Types, and Antimicrobial Resistance in *Vibrio parahaemolyticus* and *Vibrio vulnificus*: A Whole Genome Sequencing Study of Clinical and Seafood Isolates in New Jersey

**DOI:** 10.64898/2026.07.30.26359351

**Authors:** L Schlitt, FM Wu, B Jeong, L Bodnar, A Panyi, J Bilinski, M Steinberg, C. Lloyd, J. Hutcheson, A Oyelade, M Carayannopoulos, T Kirn, F Nindo

**Author notes:** **Corresponding authors:** Lisa A. Schlitt, Fredrick Nindo.

## Abstract

*Vibrio parahaemolyticus* and *Vibrio vulnificus* are significant foodborne pathogens linked to seafood consumption and environmental exposure. Whole genome sequencing (WGS) was performed on *Vibrio* isolates collected from clinical cases and seafood sources throughout the state of New Jersey to elucidate genomic diversity, antimicrobial resistance (AMR) profiles. Sequences were included from isolates collected over eight years, from June 2016 to November 2024. This study identified 465 *Vibrio* sequences, 406 sequences from seafood sources and 59 sequences from clinical cases. Species-level taxonomic identification via Kraken2 classified 300 isolates as *Vibrio parahaemolyticus* and 165 as *Vibrio vulnificus* from whole genome assemblies. Multi-locus sequence typing (MLST) indicated a diverse population of isolates, with 169 known *Vibrio* sequence types (STs) identified. An additional 168 potential novel allelic profiles were identified, comprising 19.3% of *V. parahaemolyticus* sequences and 90.9% of *V. vulnificus* sequences. Novel sequence types were submitted to pubMLST for classification, resulting in the identification of 49 novel *V. parahaemolyticus* STs and 113 novel *V. vulnificus* STs. Three *V. parahaemolyticus* sequence types were identified in both clinical and environmental sequences. A single novel sequence type was identified in both clinical and environmental sequences of *V. vulnificus*. Analysis of antimicrobial resistance genes revealed the presence of the tetracycline resistance gene *tet(34)* in nearly all isolates. Beta-lactamase genes were detected in nearly all *V. parahaemolyticus* sequences but were absent from *V. vulnificus*, with gene profiles varying by sequence type. To the best of our knowledge, this study provides the first comprehensive WGS-based analysis of the genomic diversity and antimicrobial resistance profiles of *Vibrio parahaemolyticus* and *Vibrio vulnificus* isolates from clinical and seafood sources in New Jersey over an eight-year period, to support public health surveillance of these foodborne pathogens.

## Introduction

The genus *Vibrio* comprises over 100 known species of aquatic bacteria, 12 species of which pose serious public health concerns due to their ability to cause gastrointestinal and wound infections in humans [1]. *Vibrio parahaemolyticus* and *Vibrio vulnificus* are two infectious species of critical importance. These bacteria are commonly found in marine and estuarine environments and can cause foodborne illnesses through the consumption of contaminated seafood, particularly raw or undercooked oysters [1]. As climate change continues to warm coastal waters, *Vibrio* infections are projected to increase in New Jersey [2], making robust surveillance systems more crucial than ever.

Whole genome sequencing (WGS) has revolutionized microbial genomics and public health surveillance by enabling high-resolution tracking of pathogen evolution, transmission, and resistance mechanisms. We hypothesized that V. parahaemolyticus and V. vulnificus isolates would show distinct sequence type (ST) and antimicrobial (AMR) profiles separating seafood and clinical isolates; ST and AMR profiles identified in both clinical and environmental sequences would indicate a possible transmission event. Phylogenomic analysis of these sequences could then be used to uncover evolutionary links and transmission dynamics from emerging high-risk lineages, enabling WGS surveillance for seafood safety. This study leverages WGS to investigate the genomic epidemiology of V. parahaemolyticus and V. vulnificus isolated from clinical and environmental (seafood-oyster) surveillance samples. Our goals were to predict the biotypes [sequence types (STs)], characterize antimicrobial resistance (AMR) profiles, and generate recombination-aware SNP-based phylogenomic trees that improves our molecular characterization of pathogens of public health importance. By integrating these analyses, we aimed to gain insights into strain diversity, transmission dynamics, and the emergence of antimicrobial resistance traits; information that could contribute to formulation of improved food safety policies and public health interventions.

## Materials and Methods

### Sample Collection and DNA Sequencing

Analyses of suspected *Vibrio parahaemolyticus* and *Vibrio vulnificus* isolates are regularly performed as part of New Jersey’s contribution to the CDC PulseNET program [3]. This study examined sequence reads from all 465 *Vibrio* sequences previously identified as either *V. parahaemolyticus* or *V. vulnificus* over an eight-year period spanning from 2016 to 2024.

Clinical isolates of *V. parahaemolyticus* and *V. vulnificus* were obtained through routine case-based surveillance from patients presenting with gastrointestinal or wound infections in medical centers participating in foodborne pathogen surveillance program around the state of New Jersey. As a tertiary state public health reference laboratory engaged in federally supported surveillance, outbreak detection, and response for infectious diseases, the New Jersey Public Health and Environmental Laboratories routinely receive pure culture isolates from submitting clinical, hospital, and local diagnostic laboratories for confirmatory testing and genomic characterization.

Seafood isolates were collected from shellfish samples during targeted surveillance of nine commercial harvesting sites along Delaware Bay at the southern edge of New Jersey. Samples were collected over an eight-year span, from June 2016 to November 2024. Eastern oyster (*Crassostrea virginica*) and hard clam (*Mercenaria mercenaria*) were randomly selected from commercial harvests for analysis. Each sample consisted of approximately 12-15 animals homogenized in phosphate buffered saline. *Vibrio* colonies were identified using chromogenic culture media.

Genomic DNA from *Vibrio parahaemolyticus* and *Vibrio vulnificus* isolates was extracted following the CDC PulseNet [3] standardized protocol for bacterial DNA extraction. DNA libraries were prepared using the Illumina DNA Prep Kit (Illumina, San Diego, CA) according to the manufacturer’s instructions and PulseNet SOPs. Whole genome sequencing was performed on the Illumina MiSeq platform using paired-end chemistry.

### Post-sequencing Analysis and Genome Assembly

Raw sequencing reads were trimmed using fastp as paired reads with default parameters [4]. Trimmed sequences were assessed for quality with the FastQC package [5]. Sequences were selected for assembly if at least one read for the pair had a median per base quality score of 25 or higher, and a lower quartile score greater than 10.

Prior to assembly, sequence reads were assigned taxonomic classifications using the Kraken2 Standard- 8 database [6]. Reads were filtered for *Vibrio* sequences using KrakenTools [7]; this increased the aligned genome fraction percentage while reducing the number of misassembles. *De novo* genome assembly was performed using SPAdes [8]. Assembly quality for each species was assessed by aligning contigs to the NCBI Reference Genome using Quast 5.3 [9].

### Multi-Locus Sequence Typing (MLST)

Assembled scaffolds were assigned sequence types by querying them against the PubMLST database [10]. *Vibrio parahaemolyticus* sequence typing uses a profile of seven genes as described by Gonzalez- Escalona *et al.* [11]. The typing scheme for *Vibrio vulnificus* classifies isolates according to ten loci, five from each chromosome, as originally described by Bisharat *et al.* [12].

Novel sequence types were assigned automatically if an existing allele profile was not identified in the PubMLST database. Potential novel alleles were identified using the MLST tool developed by Seemann [13], including both full length novel alleles and partial matches. Alleles were classified as novel variants if there was no exact match in the PubMLST database; all novel alleles were submitted to PubMLST for review and classification. Alelle profiles with no match to an existing sequence type were submitted to PubMLST for sequence type assignment.

### Preliminary Phylogenomic Analysis

Assemblies were annotated to predict/identify the presence of antimicrobial resistance (AMR) and virulence genes in each sequence. Antimicrobial resistance genes were identified using AMRFinderPlus [14].

Whole genomes were aligned using Snippy v4.60 (https://github.com/tseemann/snippy) for each species [15]. Phylogenetic trees were estimated from whole genome alignments using FastTree 2.1 with the Jukes-Cantor model and 1,000 iterations [16].

Phylogenetic trees were annotated using FigTree v.1.44 (https://github.com/rambaut/figtree). Heatmap diagrams were created using the ggtree R package [17].

### Ancestral Reconstruction

To evaluate the *V. parahaemolyticus* habitat switch from environment to human or vice versa, mixed clades (containing sequences from both seafood and clinical samples) obtained from maximum likelihood tree obtained from prior phylogeny were selected for further analysis. Core SNP alignments were constructed using Snippy v4.60; this alignment was then filtered using Gubbins v3.4.1 to account for recombinant regions [18].

The core SNP alignment was analyzed using TempEST v1.5.3 [19] to confirm the presence of molecular clock-like behavior. BEAST2 v2.7.7 [20] was used for phylogenetic reconstruction of ancestral sources using a Coalescent Bayesian Skyline tree prior. A Markov chain Monte Carlo analysis was run for 500 million states to ensure adequate mixing and convergence. A general time reversible (GTR) substitution model and a relaxed log normal clock model were applied based on the inference from Jmodeltest [21, 22].

## Results

A total of *465 Vibrio* sequences were successfully taxonomically identified and assembled for downstream analysis. *Vibrio parahaemolyticus* was the most abundant species (n=300 isolates) representing 64.5% of sequences, with *Vibrio vulnificus* (n=165) comprising 35.5% of the classified sequences. Most isolates, 406 out of 465 (87.3%) were sampled from raw seafood sources. The remaining 59 sequences (12.7%) originated from human clinical specimens (Table 1).

**Table 1:** Identified Vibrio species by isolate source.

| Species | Number Identified | Source |  |
| --- | --- | --- | --- |
|  |  | Seafood (87.3%) | Clinical (12.7%) |
| *Vibrio parahaemolyticus* | 300 (64.5%) | 254 | 46 |
| *Vibrio vulnificus* | 165 (35.5%) | 152 | 13 |
| **Totals** | 465 | 406 | 59 |

### Vibrio parahaemolyticus Sequence Analysis

#### Multilocus Sequence Typing (MLST)

*Vibrio parahaemolyticus* sequences represented a diverse population of isolates. A total of 159 unique sequence types (STs) were identified. The most common ST identified was ST36, which was identified in 12 sequences, all sourced from clinical specimens. Most STs were unique; 121 MLSTs were only identified in a single sequence.

Fifty-two sequences (17.3%) contained allele profiles that could not be matched to an existing sequence type in the PubMLST database. A total of 49 novel sequence types were identified and assigned to novel identifiers in the PubMLST database (Supplemental Table 1). Forty-six novel sequence types corresponded only to a single isolate each; the remaining three sequence types (ST4292, ST4293, ST4311) were each identified in two separate isolates. No novel sequence types were shared by seafood and clinical isolates.

An examination of the seven sequence type alleles identified 756 known alleles, and 46 novel alleles from the 46 novel sequence type profiles. A total of 41 novel alleles were identified in sequences from seafood, with the remaining 5 novel alleles identified in sequences from clinical isolates (Table 2).

**Table 2:** ***Vibrio parahaemolyticus* sequence type alleles**: Identified and novel alleles of seven housekeeping genes used in the sequence typing scheme by Gonzalez-Escalona et al (2008)

| Gene | All Samples |  | Human |  | Seafood |  |
| --- | --- | --- | --- | --- | --- | --- |
|  | Known | Novel | Known | Novel | Known | Novel |
| *dnaE* | 107 | 2 | 20 | 0 | 99 | 2 |
| *dtdS* | 120 | 6 | 22 | 0 | 108 | 6 |
| *gyrB* | 133 | 9 | 23 | 0 | 118 | 9 |
| *pntA* | 86 | 3 | 18 | 1 | 80 | 2 |
| *pyrC* | 119 | 14 | 17 | 3 | 111 | 11 |
| *recA* | 112 | 6 | 22 | 0 | 103 | 6 |
| *tnaA* | 79 | 6 | 18 | 1 | 74 | 5 |
| **Total** | 756 | 46 | 140 | 5 | 693 | 41 |

#### Phylogenetic Analysis

Phylogenetic analysis largely indicated relationships based on identified sequence type. A tree of all assembled sequences was constructed, depicting human lineages in red and seafood lineages in blue (Fig. 1). Only three sequence types were identified in both human and seafood isolates: ST3, ST32, and ST3570. With limited metadata available, no definitive crossover events between human and seafood isolates could be identified. Most identified sequence types were limited to either human or seafood isolates A notable example of the separation between human and seafood isolates is visible in the middle of the tree, depicting a large human lineage containing sequence types ST36 and ST636.

**Fig 1:**
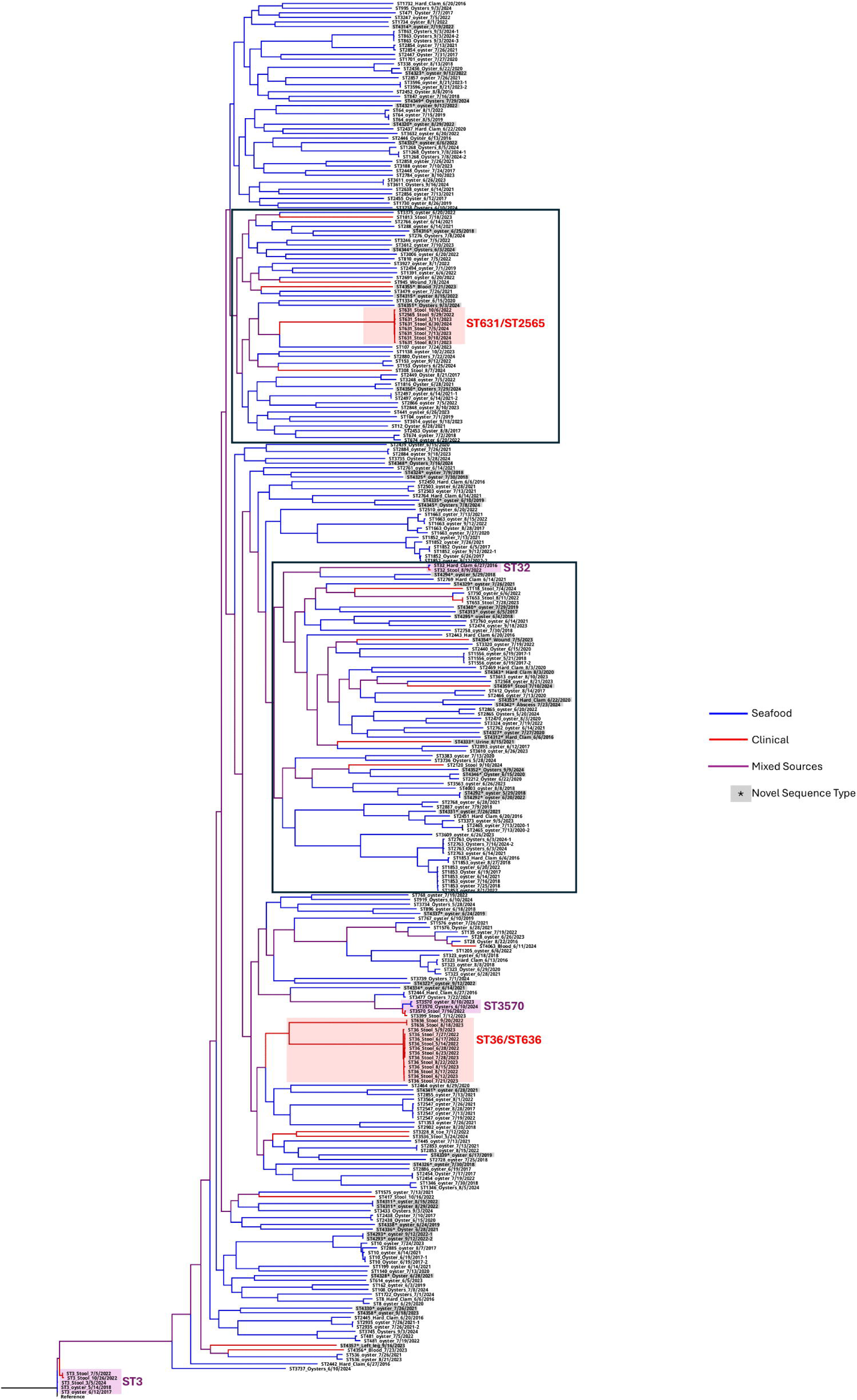
Whole genome phylogeny of 300 *Vibrio parahaemolyticus* sequences identified in New Jersey. Lineages found in seafood are colored blue and in clinical specimens red. A purple color indicates a lineage containing both seafood and human sequences. Novel sequence types identified in this study are highlighted in gray and denoted by an asterisk (*). Outlined clades include sequences selected for ancestral reconstruction.

#### AMR profiles

Analysis of the assembled genomes using AMRFinderPlus revealed 19 antimicrobial profiles; all 300 sequences were found to contain at least three genes associated with antimicrobial resistance. The most common profile (*blaCARB-18, tet(34), tet(35)*) was identified in 117 sequences and was present in both seafood and human isolates. The *tet(34)* gene, which encodes a magnesium dependent tetracycline resistance mechanism [18], was identified in all *V. parahaemolyticus* sequences. (Fig. 2).

**Fig 2.**
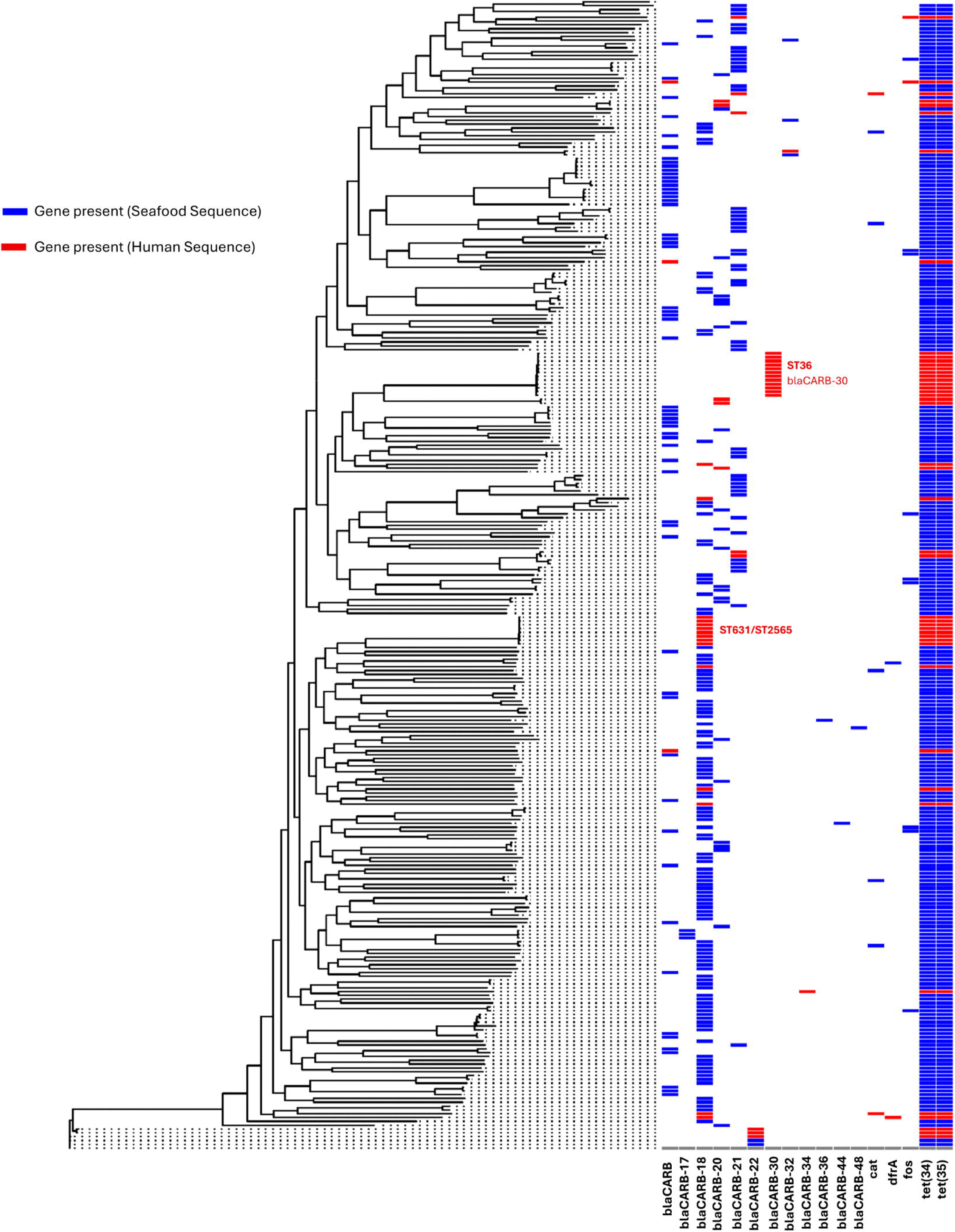
Vibrio parahaemolyticus AMR profiles. AMR profiles mapped to the phylogenetic tree of all *Vibrio parahaemolyticus* isolates; seafood sequences are shown in blue and human sequences in red. The large bands showing the common tetracycline resistance genes (*tet34* and *tet35)* are immediately noticeable. Also notable is the ST36 cluster of human isolates, containing the blaCARB-30 gene.

#### Ancestral State Reconstruction

Three mixed clades containing sequences from both seafood and clinical sources were selected for Bayesian reconstruction of the ancestral states. A core alignment of 183 sequences was created using Snippy, consisting of 456 sites. Prior to ancestral state phylogenetic inference, temporal signal was assessed and confirmed using root-to-tip regression analysis in Tempest, demonstrating a meaningful correlation between genetic divergence and sampling time. This validation supported the application of a molecular clock framework and justified the use of time-scaled phylogenetic reconstruction. Consequently, the resulting phylogeny enabled not only the resolution of evolutionary relationships but also the inference of probable directional transitions between ecological states over time.

The tree in Fig 3 depicts the estimated source of each ancestral node. These results depict the seafood source state, in blue, as the ancestral state for all tested clades. The phylogenetic structure consistently indicated a seafood-associated origin, even when contemporary isolates were derived from both clinical and environmental sources. Analysis was conducted using a compact yet information-dense whole genome SNP alignment comprising 183 sequences and 456 variable sites, specifically selected to capture vertically inherited evolutionary signals while minimizing confounding effects from accessory genome variation and horizontal gene transfer. This methodological choice enhanced the robustness of inferred clustering patterns and strengthened confidence in downstream ancestral state reconstruction.

**Fig 3:**
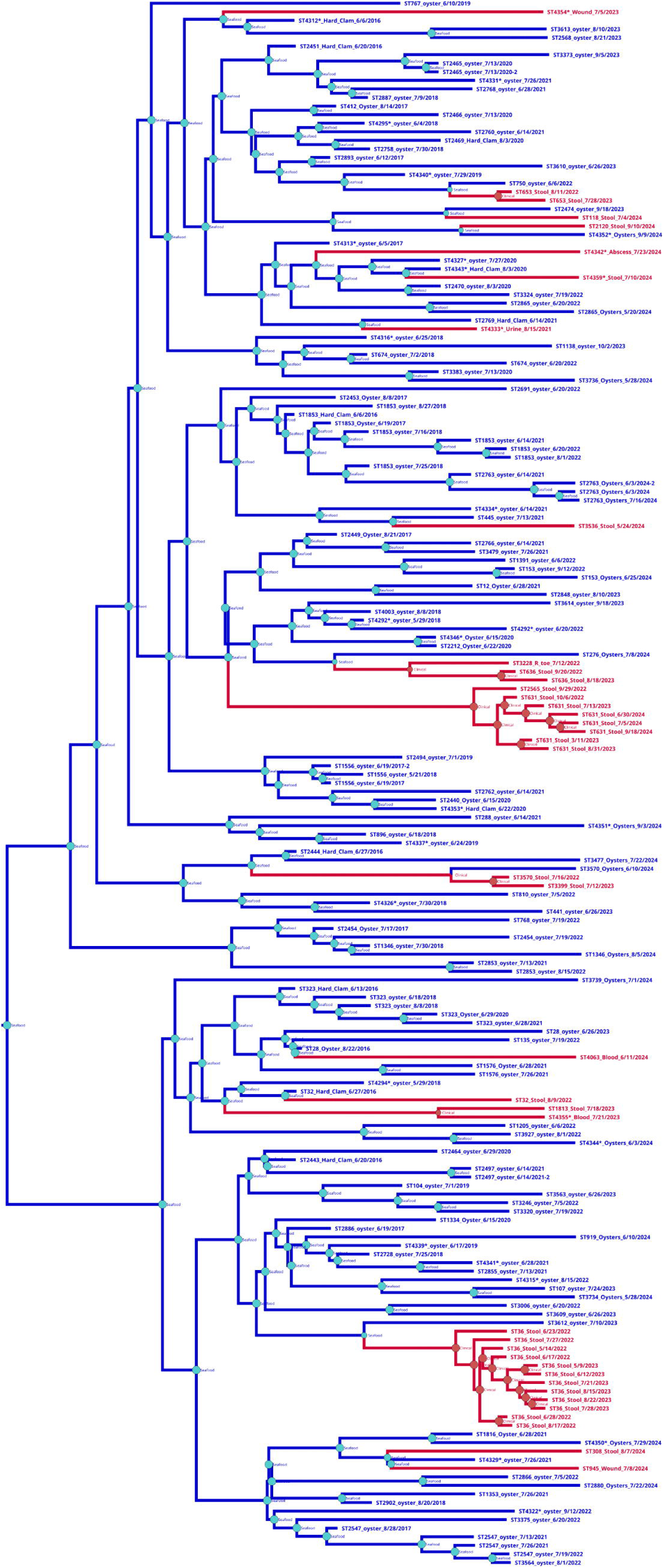
Ancestral reconstruction of mixed *Vibrio parahaemolyticus* clades containing both Seafood and Clinical sequences. Lineages found in seafood are colored blue and in clinical specimens red. Reconstruction suggests all sequences originate from seafood ancestral populations.

### Vibrio vulnificus Sequence Analysis

#### Multilocus Sequence Typing (MLST)

*Vibrio vulnificus* isolates largely contained novel allelic profiles, with 150 sequences (90.9%) that could not be matched to an existing sequence type record in PubMLST. Only ten known sequence types were identified, with the most common sequence type, ST253, only detected in five isolates.

Sequences with unknown sequence types were submitted to the PubMLST database for sequence type assignment. This study identified 113 novel sequence types (Supplemental Table 2). The most frequent novel sequence type, ST780, was identified in five seafood isolates. One novel sequence type, ST799, was identified in both a seafood and a clinical sequence; no epidemiological link between these samples was known.

Analysis of the ten alleles used in the *V. vulnificus* typing scheme identified 281 known allele variants and 101 novel alleles (Table 3). Ninety-one novel alleles were exclusively detected in seafood-derived sequences, with only 10 novel alleles identified in clinical isolates. No novel alleles were present in ST799, the novel crossover sequence.

**Table 3:**
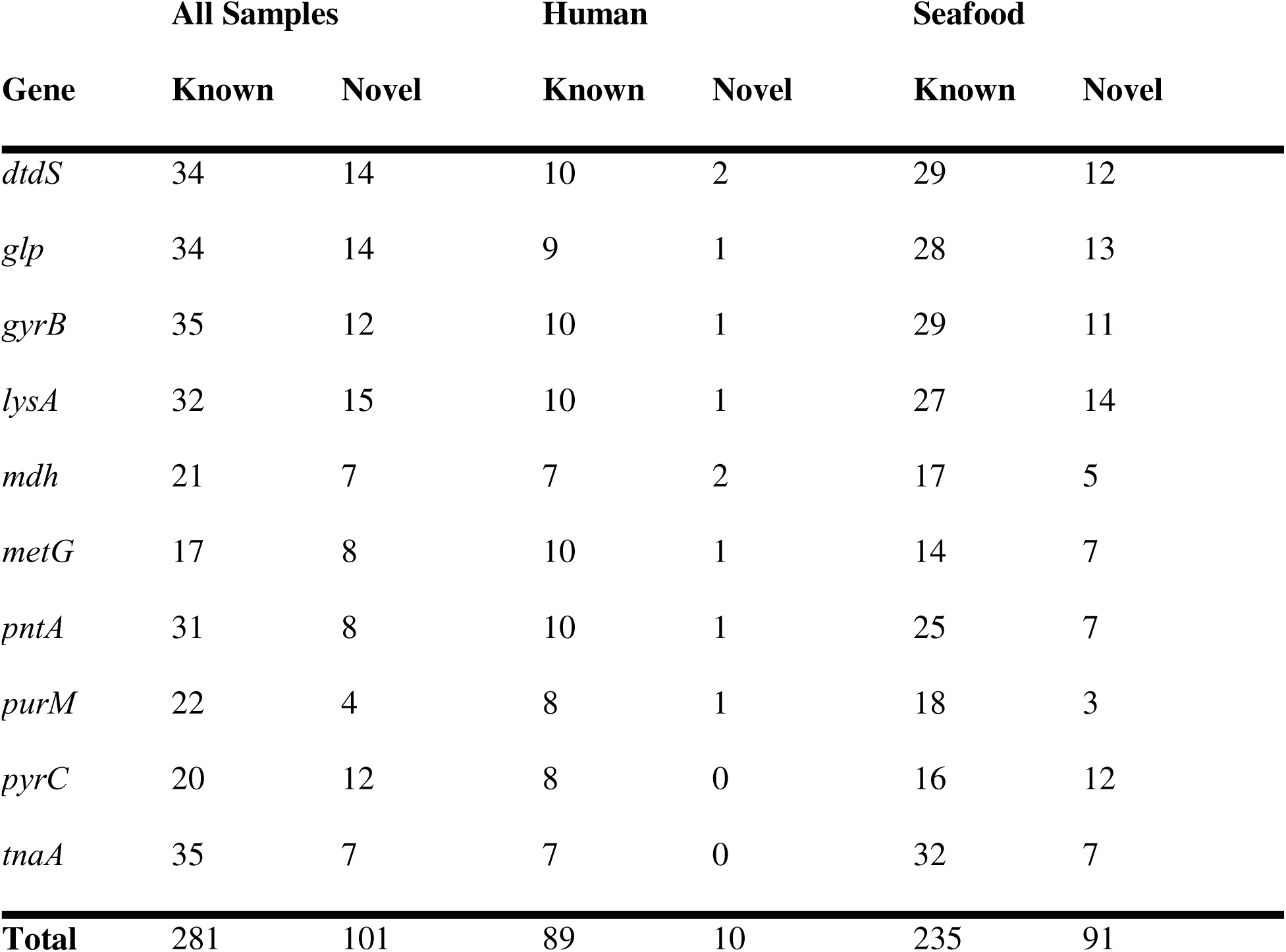
***Vibrio vulnificus* sequence type alleles:** Identified and novel alleles ten housekeeping genes used in the sequence typing scheme by Bisharat et al (2007)

| Gene | All Samples |  | Human |  | Seafood |  |
| --- | --- | --- | --- | --- | --- | --- |
|  | Known | Novel | Known | Novel | Known | Novel |
| *dtdS* | 34 | 14 | 10 | 2 | 29 | 12 |
| *glp* | 34 | 14 | 9 | 1 | 28 | 13 |
| *gyrB* | 35 | 12 | 10 | 1 | 29 | 11 |
| *lysA* | 32 | 15 | 10 | 1 | 27 | 14 |
| *mdh* | 21 | 7 | 7 | 2 | 17 | 5 |
| *metG* | 17 | 8 | 10 | 1 | 14 | 7 |
| *pntA* | 31 | 8 | 10 | 1 | 25 | 7 |
| *purM* | 22 | 4 | 8 | 1 | 18 | 3 |
| *pyrC* | 20 | 12 | 8 | 0 | 16 | 12 |
| *tnaA* | 35 | 7 | 7 | 0 | 32 | 7 |
| **Total** | 281 | 101 | 89 | 10 | 235 | 91 |

#### Phylogenetic Analysis

A phylogenetic tree of all *Vibrio vulnificus* (Fig 4) isolates shows a greater separation between seafood and clinical isolates, with a large clade of sequences only found in seafood visible in the middle of the tree. Immediately notable is the distant clade containing the majority of sequences associated with human specimens, suggesting these isolates are not common in the larger *V. vulnificus* population. The remaining clinical sequences are dispersed throughout the tree, appearing to share closer ancestry with the other seafood sequences.

**Fig 4:**
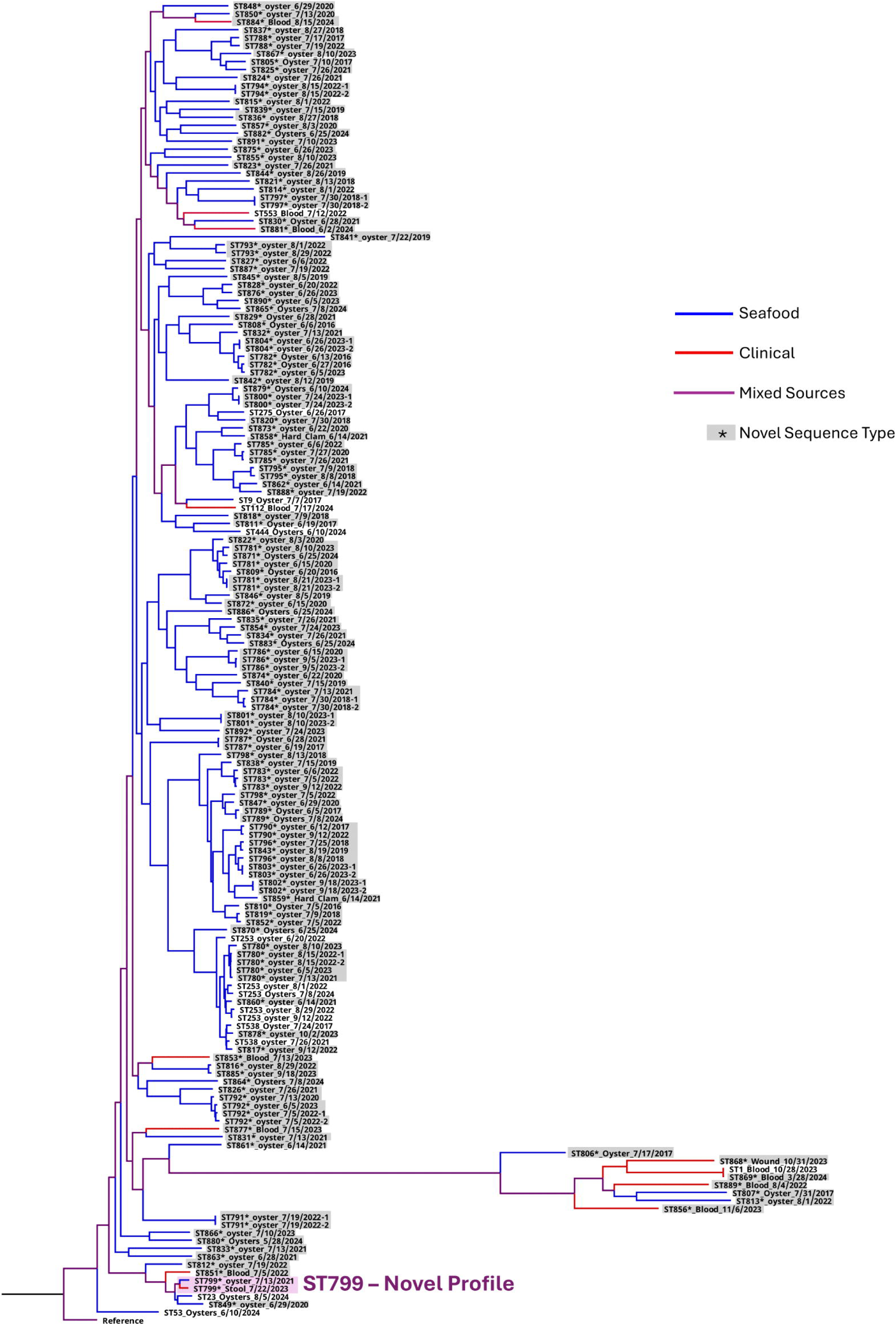
Whole genome phylogeny of 165 *Vibrio vulnificus* sequences identified in New Jersey. Lineages found in seafood are colored blue, and from human clinical samples, red. A purple color indicates a lineage containing both seafood and human sequences. Novel sequence types identified in this study are highlighted in gray and denoted by an asterisk (*).

*V. vulnificus* isolates from New Jersey did not show significant variation in their predicted antimicrobial profiles, with 163 of 165 isolates containing only the *tet(34)* tetracycline resistance gene. A single sequence from a seafood sample (oyster) contained a fosfomycin resistance gene (*fos*). These genes are known to be plasmid associated, suggesting the possible presence of a plasmid in this isolate. No beta lactamase genes were identified in this species. No antimicrobial genes were identified in a single sequence isolated from a clinical case (Fig 5).

**Fig 5.**
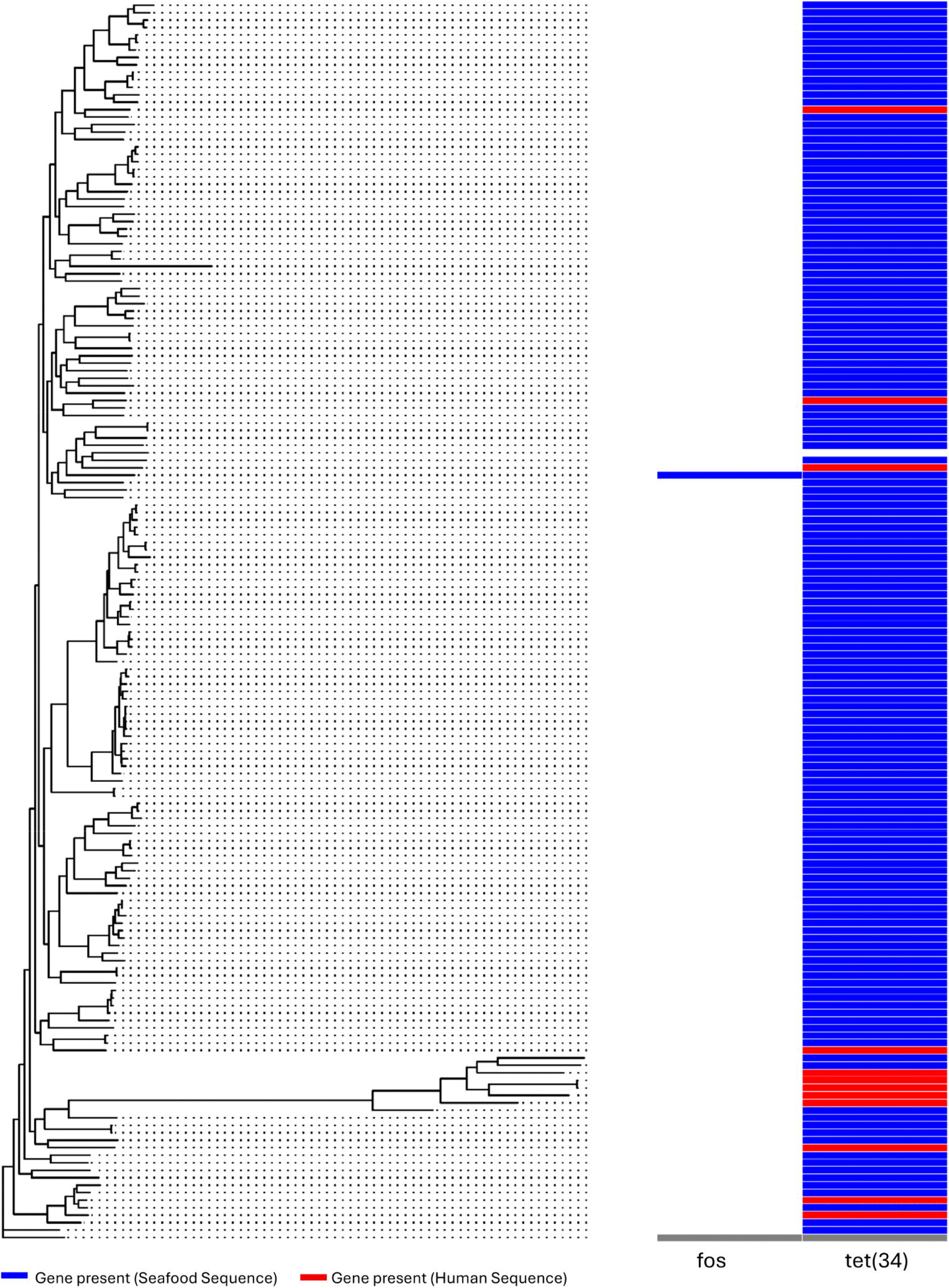
*Vibrio vulnificus* AMR profiles. AMR profiles mapped to the phylogenetic tree of all *Vibrio vulnificus* isolates. Sequences from seafood sources are shown in blue and human sequences in red. As with *V. parahaemolyticus*, the common tetracycline resistance gene is immediately noticeable.

#### Ancestral State Analysis

We additionally sought to understand the dissemination of *V. vulnificus* between seafood and clinical sources. To this end, we identified mixed clades from our previously reconstructed maximum likelihood phylogeny. Eighteen sequences representing mixed seafood/clinical sources were selected for ancestral state reconstruction. However, an examination of the core SNP alignment using TempEST did not indicate molecular clock-like behavior; based on this finding no further analysis was performed.

## Discussion

### Diversity of Sequence Types

Sequence Type (ST) analysis revealed substantial genetic diversity among both *V. parahaemolyticus* and *V. vulnificus* isolates. A total of 159 STs were identified for *V. parahaemolyticus;* although only ten known STs were identified in *V. vulnificus,* with most sequences representing possibly novel allelic profiles. Three *V. parahaemolyticus* STs, ST3, ST32, and ST3570, were identified in both clinical and seafood isolates, suggesting the possibility of intermixing of human and shellfish reservoirs for pathogenic strains.

The most frequently identified *Vibrio parahaemolyticus* sequence type was ST36, detected in 12 clinical isolates. ST36 is the most common sequence type isolated in the United States [24]. The strain is native to the Pacific Northwest, with isolates detected in the Northeast region since 2012 [25, 26]. ST36 has also spread worldwide, with isolates detected in Spain, Peru, and New Zealand [25, 26]. This sequence type is associated with a lower infective dose [26] and was the most common clinical sequence type identified in this study.

ST631, detected in seven sequences, was also notable among the clinical isolates. This strain originated in Louisiana in 2007, with the earliest New Jersey sequence (not included in this study) detected in 2013. This sequence type is known to share a similar virulence profile to ST36 and is the second most common source of clinical *V. parahaemolyticus* infections in the United States [24].

The most common sequence type worldwide, ST3 [24], was among the three *V. parahaemolyticus* STs detected in both clinical and seafood sourced sequences. This sequence type originated in India in 1996, spreading clonally to South and Central America [27]. ST3, along with ST36, is a known “pandemic” strain associated with global epidemics [28]. The presence of this sequence type in both seafood and clinical isolates suggests a potential reservoir in New Jersey shellfish warranting further investigation.

This study identified 162 novel *Vibrio* sequence types, with 49 novel sequence types identified in *V. parahaemolyticus,* and 113 novel sequence types identified in *V. vulnificus*. A single novel sequence type, from *V. vulnificus*, ST799, was identified in both a seafood isolate from 2021 and a clinical specimen from 2023. The presence of this shared sequence type further supports the possibility of shellfish reservoirs for infectious strains and indicates that continual monitoring of the *Vibrio* population in New Jersey may be warranted. The current PulseNet surveillance protocols in New Jersey do not classify novel sequence types; the potential link between these sequences would not have been identified outside of this whole genome study.

### Antimicrobial resistance

This investigation into the antimicrobial resistance (AMR) profiles of *Vibrio parahaemolyticus* and *Vibrio vulnificus* revealed the presence of shared genes across all isolates regardless of source. The tetracycline resistance genes *tet(34)* and *tet(35)* were detected in all 300 *V. parahaemolyticus* sequences. The *tet(34)* gene was also detected in all but one *V. vulnificus* isolate.

Additional antimicrobial genes in *V. parahaemolyticus* isolates varied largely according to sequence type. The beta lactamase *blaCARB-30* was detected only in ST36 isolates, *blaCARB-22* in ST3, and *blaCARB-17* in ST863. The most common profile (*blaCARB-18, tet(34), tet(35)*) was detected in 117 isolates comprising 72 unique sequence types.

These findings suggest that the marine environment, particularly aquaculture systems, may be a potential reservoir or conduit for AMR dissemination [29]. Oxytetracycline is a common antibiotic used in fisheries and aquaculture [23, 30]. A previous study in China also identified the tet(34) and tet(35) resistance genes across all *V. parahaemolyticus* isolates; however only 3% of these isolates showed tetracycline resistance using phenotypic screening methods[31]. As antimicrobial susceptibility testing is not currently performed as part of the existing surveillance program in New Jersey, the extent of antimicrobial resistance in the Vibrio population remains unknown.

This data underscores the urgent need for ongoing AMR monitoring in marine environments, not only to ensure food safety but also to better understand the broader dynamics of resistance gene spread, which could inform strategies to mitigate the growing threat of AMR [32].

### Ancestral State Reconstruction

Across all three mixed clades analyzed, ancestral state reconstruction consistently resolved basal nodes to a seafood-associated origin. This finding was notable given that these clades contained interspersed clinical and environmental isolates. Under a model of independent clinical adaptation or sustained human transmission, one would expect at least a subset of ancestral nodes to resolve to a clinical state. However, the persistent inference of seafood-associated ancestry suggested that clinical isolates arose from repeated spillover events originating in marine or seafood reservoirs rather than forming stable, human-adapted lineages.

From an evolutionary perspective, this pattern was consistent with an environmentally maintained pathogen exhibiting zoonotic-like dynamics. Clinical isolates appeared predominantly as terminal or recently derived branches, supporting the interpretation of recent host transitions rather than long-term persistence in human populations. The Bayesian framework further reinforced this conclusion, as ancestral state probabilities incorporated both phylogenetic topology and branch length information, indicating a strong and consistent signal across the dataset.

These findings carried important implications for public health surveillance. The dominance of seafood- associated ancestral states suggested that clinical surveillance alone captured only a limited representation of the *V. parahaemolyticus* genetic diversity. The majority of evolutionary diversification appeared to occur within marine environments, aquaculture systems, or perhaps along seafood supply chains, with human infections representing downstream events. As such, reliance solely on clinical isolates would likely underestimate both pathogen diversity and the emergence of novel lineages.

## Conclusions

This genomic epidemiology surveillance investigation examined two clinically and environmentally significant bacterial species, *Vibrio parahaemolyticus* and *Vibrio vulnificus*, isolated from both clinical and shellfish sources. Through whole genome sequencing (WGS) and advanced bioinformatics analyses, we characterized the population structure, antimicrobial resistance (AMR) profiles, and transmission dynamics of these pathogens. Our results provide critical insights into pathogen dissemination patterns and have direct implications for food safety surveillance and public health policy.

The application of WGS enabled comprehensive testing of hypotheses regarding bacterial adaptation mechanisms across environmental gradients, antimicrobial resistance acquisition pathways, and potential epidemiological linkages between clinical and environmental reservoirs. The detection of shared sequence types and antimicrobial profiles in some environmental and clinical isolates highlights the role of coastal/estuarine and marine ecosystems as probable reservoirs of pathogenic *Vibrio* species. This may have direct implications for seafood safety, especially in the context of raw oyster consumption. Public health agencies can leverage these findings to guide formulation of targeted advisories and prioritize monitoring of specific STs or virulence signatures in that could increase risk of Vibrio associated infections.

Analysis of novel sequence types led to the discovery of a novel *Vibrio vulnificus* sequence type, ST799, present in both seafood and clinical samples. Current surveillance practices in New Jersey limit sequence type identification to known records in the pubMLST database; novel sequence types are noted but unclassified. The identification of ST799 demonstrates the potential for whole genome sequencing to identify linked specimens with novel allele profiles that may otherwise be overlooked during normal surveillance. It may be prudent to integrate the classification of novel sequence types as part of New Jersey’s regular sequencing protocols.

The results may also have implications for outbreak investigation. The embedding of clinical isolates within broader seafood-associated diversity indicated that genomic similarity among patient isolates might reflect shared environmental exposure rather than direct human-to-human transmission. This observation complicates traditional epidemiological interpretations and highlighted the necessity of integrating genomic data with food traceability and environmental sampling frameworks.

From a risk assessment standpoint, the consistent inference of seafood-associated ancestry suggested that adaptation to the human host remained incomplete or transient. While this may limit sustained transmission within human populations, it simultaneously underscored the role of the environmental reservoir as a continuous source of genetic innovation. This reservoir has the potential to generate strains with enhanced virulence or antimicrobial resistance, emphasizing the importance of proactive genomic surveillance in non-clinical settings.

These findings underscore the necessity of integrating WGS into routine surveillance frameworks. Climate driven ocean warming and increased globalization of food distribution networks have the potential of contributing to the geographic expansion and increased prevalence of *Vibrio* species. Elevated water temperatures may enhance bacterial proliferation in marine ecosystems, while international trade could facilitate pathogen introduction into previously unaffected regions. The convergence of environmental and clinical transmission pathways presents significant challenges for disease control efforts. Implementation of genomic surveillance systems is therefore essential for early detection and rapid response to emerging threats, thereby contributing to the reduction of the burden of *Vibrio*-associated foodborne illnesses. We however wish to reiterate that in general surveillance studies such as the study at hand has several limitations but more importantly surveillance data, while it may be geographically comprehensive, relies on whoever seeks care or gets tested— forming a convenience sample skewed by access, awareness, and behavior. This non-random collection may miss asymptomatic cases, underserved populations, and those avoiding medical systems, thereby creating blind spots that distort true disease patterns and burden estimates.

## Future Directions

Several research priorities emerge from this investigation. First, expansion of WGS-based surveillance programs should encompass a broader taxonomic range of seafood vectors (including crustaceans and finfishes) and diverse aquatic environments (rivers, estuaries, and aquaculture operations). This comprehensive sampling strategy would facilitate identification of high-risk ecological niches and enable targeted interventions earlier in the food production chain.

Second, longitudinal genomic surveillance incorporating temporal sampling across multiple years and seasonal cycles is warranted. Such analyses would elucidate evolutionary dynamics, environmental adaptation mechanisms, and antimicrobial resistance emergence patterns in response to climatic variability and anthropogenic pressures. Time-series genomic data would enhance predictive modeling capabilities for outbreak risk assessment and support evidence-based resource allocation for prevention efforts.

This study also identified areas for improved collaboration across disparate agencies. Specimens included for analysis were collected routinely as part of environmental and clinical surveillance programs; sequencing of isolates was performed at a later date. As a result, metadata was difficult to reconcile with sequences as identifiers varied at each stage of analysis. As sequencing programs are expected to play a larger role in public health, the possibility of downstream analysis needs to be considered at earlier stages of future analyses.

Delving into metagenomic studies of oyster microbiomes offers exciting potential to uncover the broader microbial communities within these shellfish. Such investigations could detect not only co- occurring pathogens but also the collective resistomes—the array of antibiotic resistance genes present—shedding light on interactions that might amplify threats to human health. Understanding these complex ecosystems at a molecular level could lead to innovative strategies for mitigating risks, such as developing probiotics or targeted treatments to disrupt harmful bacterial networks.

Finally, the development of integrated risk models stands out as a critical step forward. These models would combine genomic data with environmental factors, like water quality and climate metrics, alongside epidemiological insights from human cases.

## Statements

### Statement of Ethics

Data for this study was collected through normal public health surveillance; ethical approval is not required for this study in accordance with guidelines set by the Rowan University IRB acting on behalf of the New Jersey Department of Health.

The opinions expressed in this manuscript are the author’s own and do not reflect the view of the New Jersey Department of Health, the State of New Jersey, or the United States Food and Drug Administration.

### Author Contributions

LS performed all included analyses under the direction of FN and prepared all figures and tables. CL and JH collected and prepared seafood samples. Sequencing was performed by FW, LB, JB, MS and AP, under the direction of AO. Initial analysis and species identification was performed by BJ. The manuscript was prepared by LS and FN, with review and support from TK and MC.

### Data Availability Statement

All analyzed sequences were submitted to the Sequence Read Archive (SRA) at the National Center for Biotechnology Information (NCBI) (https://www.ncbi.nlm.nih.gov/sra). *Vibrio parahaemolyt*icus accession numbers are listed in Supplemental Table 3 and *Vibrio vulnificus* accession numbers are listed in Supplemental Table 4.

