## Supplemental Table 1 for "Population Structure, Novel Sequence Types, and Antimicrobial Resistance in *Vibrio parahaemolyticus* and *Vibrio vulnificus*: A Whole Genome Sequencing Study of Clinical and Seafood Isolates in New Jersey"

**Supplemental Table 1**: Novel *Vibrio parahaemolyticu*s Profiles. Alleles designated by italics and an asterisk (*) represent novel alleles identified as part of this study.

| MLST Profile Gene | | | | | | | Assigned ST | Sequences | Sources |
| --- | --- | --- | --- | --- | --- | --- | --- | --- | --- |
| *dnaE* | *dtdS* | *gyrB* | *pntA* | *pyrC* | *recA* | *tnaA* |  |  |  |
| 26 | 105 | 57 | 5 | *656** | 200 | *424** | ST4292 | 2 | Seafood (Oyster) |
| 7 | 26 | 236 | 6 | 18 | 4 | 26 | ST4293 | 2 | Seafood (Oyster) |
| 20 | 27 | 16 | 42 | 29 | 15 | 23 | ST4294 | 2 | Seafood (Oyster) |
| 20 | *724** | 61 | 206 | 29 | 6 | *425** | ST4295 | 1 | Seafood (Oyster) |
| 3 | 75 | 101 | *396** | 11 | *655** | 428* | ST4311 | 1 | Seafood (Oyster) |
| 28 | 36 | *774** | 33 | 356 | *650** | 20 | ST4312 | 1 | Seafood (Clam) |
| 45 | 59 | 60 | 14 | *659** | 55 | 7 | ST4313 | 1 | Seafood (Oyster) |
| 10 | 421 | 595 | 31 | 11 | 57 | 375 | ST4314 | 1 | Seafood (Oyster) |
| 158 | 35 | 104 | 45 | *663** | 19 | 57 | ST4315 | 1 | Seafood (Oyster) |
| 382 | 180 | 13 | 31 | *657** | 60 | 57 | ST4316 | 1 | Seafood (Oyster) |
| 279 | 228 | 136 | 26 | 62 | 27 | 79 | ST4320 | 1 | Seafood (Oyster) |
| 26 | 88 | 482 | 128 | 11 | 354 | 23 | ST4321 | 1 | Seafood (Oyster) |
| 10 | 34 | 5 | 20 | 22 | 17 | 15 | ST4322 | 1 | Seafood (Oyster) |
| 36 | 14 | 184 | 28 | 192 | 31 | 84 | ST4323 | 1 | Seafood (Oyster) |
| 10 | 25 | 333 | 26 | 562 | 596 | 23 | ST4324 | 1 | Seafood (Oyster) |
| 158 | 8 | 739 | 370 | 3 | 607 | 20 | ST4325 | 1 | Seafood (Oyster) |
| 148 | 205 | 248 | 117 | 3 | 75 | 87 | ST4326 | 1 | Seafood (Oyster) |
| 239 | 497 | 566 | 33 | *666** | 414 | 20 | ST4327 | 1 | Seafood (Oyster) |
| 155 | 170 | *778** | 104 | 156 | 653* | 111 | ST4328 | 1 | Seafood (Oyster) |
| 45 | *728** | *777** | 14 | 497 | 139 | *426** | ST4329 | 1 | Seafood (Oyster) |
| 26 | 116 | 571 | 26 | 11 | 97 | 146 | ST4330 | 1 | Seafood (Oyster) |
| *575** | 19 | 147 | *395** | 56 | 15 | 20 | ST4331 | 1 | Seafood (Oyster) |
| 51 | 191 | 8 | 165 | 95 | 390 | 23 | ST4332 | 1 | Seafood (Oyster) |
| 25 | 282 | 322 | 33 | 32 | 13 | *427** | ST4333 | 1 | Clinical (Urine) |
| 445 | 14 | 606 | 99 | 661* | 460 | 57 | ST4334 | 1 | Seafood (Oyster) |
| *574** | *725** | *775** | 4 | 204 | *651** | 19 | ST4335 | 1 | Seafood (Oyster) |
| 81 | 191 | 90 | 26 | 177 | 70 | 169 | ST4336 | 1 | Seafood (Oyster) |
| 28 | *727** | 17 | 24 | 23 | 21 | 24 | ST4337 | 1 | Seafood (Oyster) |
| 64 | 151 | 776* | 6 | 276 | 31 | 234 | ST4338 | 1 | Seafood (Oyster) |
| 51 | *726** | 404 | 54 | *658** | *652** | 15 | ST4339 | 1 | Seafood (Oyster) |
| 3 | 273 | 699 | 14 | *662** | 51 | 44 | ST4340 | 1 | Seafood (Oyster) |
| 446 | 48 | *780** | 18 | 46 | 198 | 47 | ST4341 | 1 | Seafood (Oyster) |
| 17 | 36 | 179 | *397** | *668** | 13 | 26 | ST4342 | 1 | Clinical (Abcess) |
| 179 | 505 | 559 | 283 | *660** | 219 | 2 | ST4343 | 1 | Seafood (Clam) |
| 49 | 229 | 525 | 31 | 8 | 31 | 144 | ST4344 | 1 | Seafood (Oyster) |
| 51 | *729** | *783** | 4 | 204 | *654** | 24 | ST4345 | 1 | Seafood (Oyster) |
| 31 | 7 | 296 | 270 | *667** | 13 | 87 | ST4346 | 1 | Seafood (Oyster) |
| 17 | 191 | 300 | 48 | 120 | 231 | 24 | ST4348 | 1 | Seafood (Oyster) |
| 34 | 27 | 358 | 26 | 54 | 30 | 86 | ST4349 | 1 | Seafood (Oyster) |
| 339 | 13 | 461 | 26 | 145 | 31 | 2 | ST4350 | 1 | Seafood (Oyster) |
| 332 | 75 | *782** | 66 | 583 | 89 | 17 | ST4351 | 1 | Seafood (Oyster) |
| 31 | 7 | 66 | 33 | *669** | 341 | 94 | ST4352 | 1 | Seafood (Oyster) |
| 17 | 36 | *781** | 15 | 452 | 13 | 26 | ST4353 | 1 | Seafood (Clam) |
| 19 | 31 | 31 | 353 | *664** | 8 | 20 | ST4354 | 1 | Clinical (Wound) |
| 42 | 383 | 593 | 356 | 46 | 73 | 94 | ST4355 | 1 | Clinical (Blood) |
| 80 | 76 | 263 | 28 | 224 | 290 | 23 | ST4356 | 1 | Clinical (Blood) |
| 220 | 50 | 207 | 25 | *665** | 2 | 17 | ST4357 | 1 | Clinical (Wound) |
| 26 | 353 | 109 | 296 | 11 | 81 | *429** | ST4358 | 1 | Seafood (Oyster) |
| 57 | 130 | 327 | 33 | 11 | 338 | 20 | ST4359 | 1 | Clinical (Stool) |
