## Supplemental Table 2 for "Population Structure, Novel Sequence Types, and Antimicrobial Resistance in *Vibrio parahaemolyticus* and *Vibrio vulnificus*: A Whole Genome Sequencing Study of Clinical and Seafood Isolates in New Jersey"

**Supplemental Table 2**: Novel *Vibrio vulnificu*s Profiles. Alleles designated by italics and an asterisk (*) represent novel alleles identified as part of this study.

| MLST Profile Gene | | | | | | | | | | Assigned ST | Sequences | Sources |
| --- | --- | --- | --- | --- | --- | --- | --- | --- | --- | --- | --- | --- |
| *dtdS* | *glp* | *gyrB* | *lysA* | *mdh* | *metG* | *pntA* | *purM* | *pyrC* | *tnaA* |  |  |  |
| 34 | 52 | 8 | 60 | 11 | 23 | 21 | 13 | 5 | 52 | ST780 | 5 | Seafood (Oyster) |
| 19 | 47 | 41 | 33 | 2 | 7 | 15 | 8 | 5 | 74 | ST781 | 4 | Seafood (Oyster) |
| 19 | *166** | 8 | 110 | 13 | 7 | 22 | 89 | 159 | 44 | ST782 | 3 | Seafood (Oyster) |
| 2 | 18 | 8 | 98 | 11 | 23 | *123** | 8 | *165** | 80 | ST783 | 3 | Seafood (Oyster) |
| *255** | 69 | *142** | *228** | 11 | 23 | 16 | 8 | *166** | 30 | ST784 | 3 | Seafood (Oyster) |
| 2 | 33 | 14 | 62 | 2 | 2 | 5 | 13 | 14 | *171** | ST785 | 3 | Seafood (Oyster) |
| 34 | *178** | *142** | 17 | 13 | 22 | 16 | 40 | 5 | 49 | ST786 | 3 | Seafood (Oyster) |
| 107 | 13 | 14 | 31 | 20 | 7 | 14 | 3 | 17 | 18 | ST787 | 2 | Seafood (Oyster) |
| 106 | 8 | *137** | *226** | 2 | *131** | 109 | *109** | 15 | 78 | ST788 | 2 | Seafood (Oyster) |
| 10 | 18 | *136** | 17 | 11 | 23 | *123** | 8 | *165** | 45 | ST789 | 2 | Seafood (Oyster) |
| 10 | 47 | *136** | 98 | 11 | 23 | *123** | 8 | *165** | 45 | ST790 | 2 | Seafood (Oyster) |
| 170 | 78 | 20 | 53 | 2 | 24 | 18 | 8 | 124 | 119 | ST791 | 2 | Seafood (Oyster) |
| *250** | *179** | 42 | 178 | 2 | 7 | 76 | 9 | 14 | 30 | ST792 | 4 | Seafood (Oyster) |
| *257** | 8 | 10 | 1 | 2 | 2 | 20 | 13 | *173** | 74 | ST793 | 2 | Seafood (Oyster) |
| 100 | 7 | 68 | *232** | *154** | 60 | 108 | 65 | 17 | 83 | ST794 | 2 | Seafood (Oyster) |
| 2 | 33 | 2 | 62 | 11 | 24 | 5 | 8 | 53 | 30 | ST795 | 2 | Seafood (Oyster) |
| 10 | 18 | *136** | 110 | 11 | 23 | *123** | 8 | *165** | 45 | ST796 | 2 | Seafood (Oyster) |
| 22 | 136 | 14 | 133 | 70 | 22 | 76 | 9 | 53 | 105 | ST797 | 2 | Seafood (Oyster) |
| 10 | 18 | *136** | 98 | 11 | 23 | *123** | 8 | *165** | 45 | ST798 | 2 | Seafood (Oyster) |
| 44 | 23 | 14 | 18 | 17 | 17 | 21 | 8 | 17 | 9 | ST799 | 2 | Seafood (Oyster)  Clinical (Stool) |
| *259** | 33 | 2 | 1 | 11 | 2 | 46 | 3 | 53 | 86 | ST800 | 2 | Seafood (Oyster) |
| 68 | 50 | 26 | 6 | 10 | 23 | 5 | 9 | 17 | 39 | ST801 | 2 | Seafood (Oyster) |
| 10 | 18 | *136** | 33 | 11 | 7 | *123** | 8 | *165** | 14 | ST802 | 2 | Seafood (Oyster) |
| 10 | 18 | *136** | 110 | 11 | 23 | *123** | 13 | *165** | 45 | ST803 | 2 | Seafood (Oyster) |
| 19 | *166** | 8 | 110 | 13 | 7 | 22 | 89 | 159 | 14 | ST804 | 2 | Seafood (Oyster) |
| 5 | 8 | *137** | *221** | 2 | *131** | 109 | *109** | 9 | 78 | ST805 | 1 | Seafood (Oyster) |
| *248** | 77 | 47 | 41 | 53 | 124 | 72 | 64 | 127 | 53 | ST806 | 1 | Seafood (Oyster) |
| 75 | 61 | 1 | 76 | 33 | 1 | 4 | 44 | 60 | 13 | ST807 | 1 | Seafood (Oyster) |
| 19 | 8 | 62 | 75 | 2 | 2 | 13 | 13 | 5 | 18 | ST808 | 1 | Seafood (Oyster) |
| 19 | 47 | 41 | 33 | 2 | 7 | 15 | 8 | *165** | 74 | ST809 | 1 | Seafood (Oyster) |
| 10 | 52 | *136** | 17 | 11 | 23 | *123** | 8 | *165** | 45 | ST810 | 1 | Seafood (Oyster) |
| 164 | 45 | 38 | 17 | 13 | 2 | 70 | 3 | 5 | 87 | ST811 | 1 | Seafood (Oyster) |
| 2 | 18 | 19 | 18 | 17 | 17 | 21 | 8 | 4 | 26 | ST812 | 1 | Seafood (Oyster) |
| 62 | 1 | 3 | 3 | 32 | *136** | 105 | 2 | 52 | 7 | ST813 | 1 | Seafood (Oyster) |
| 103 | *176** | 42 | 12 | 2 | 22 | 13 | 8 | *172** | 2 | ST814 | 1 | Seafood (Oyster) |
| 13 | 45 | 28 | 231 | 2 | 9 | 25 | 65 | *170** | 30 | ST815 | 1 | Seafood (Oyster) |
| 191 | 69 | 42 | 17 | 2 | 7 | 23 | 8 | 14 | 125 | ST816 | 1 | Seafood (Oyster) |
| 34 | 52 | 28 | 60 | 11 | 23 | 21 | 8 | *167** | 18 | ST817 | 1 | Seafood (Oyster) |
| 2 | 45 | 2 | 26 | 27 | 7 | 70 | 3 | 159 | *172** | ST818 | 1 | Seafood (Oyster) |
| 111 | 18 | *136** | 98 | 11 | 23 | *123** | 8 | *165** | 45 | ST819 | 1 | Seafood (Oyster) |
| 107 | 18 | 2 | 62 | 11 | 2 | 5 | 3 | 53 | 25 | ST820 | 1 | Seafood (Oyster) |
| 110 | *170** | 2 | 12 | 2 | 3 | 20 | 62 | 5 | 83 | ST821 | 1 | Seafood (Oyster) |
| 19 | 47 | 41 | 33 | 2 | 7 | 15 | 8 | *166** | 74 | ST822 | 1 | Seafood (Oyster) |
| 22 | 49 | 40 | 1 | 44 | 2 | 5 | 8 | 5 | 50 | ST823 | 1 | Seafood (Oyster) |
| *254** | 7 | *141** | *227** | 2 | 97 | 106 | 9 | 15 | 78 | ST824 | 1 | Seafood (Oyster) |
| *253** | 8 | *137** | *226** | 2 | *131** | 109 | *109** | 9 | 78 | ST825 | 1 | Seafood (Oyster) |
| *250** | *169** | 24 | *225** | 20 | 7 | 76 | 3 | 123 | 30 | ST826 | 1 | Seafood (Oyster) |
| *256** | *174** | *143** | 110 | 13 | 22 | 22 | 8 | *170** | 39 | ST827 | 1 | Seafood (Oyster) |
| 230 | 82 | 102 | 110 | 11 | 7 | 76 | 3 | *166** | 44 | ST828 | 1 | Seafood (Oyster) |
| 19 | 47 | 62 | 57 | 13 | 24 | 13 | 13 | 5 | 129 | ST829 | 1 | Seafood (Oyster) |
| 69 | 64 | 75 | 12 | 10 | 12 | 13 | 18 | *169** | *170** | ST830 | 1 | Seafood (Oyster) |
| *251** | 29 | *140** | 17 | 19 | 7 | 5 | 8 | 17 | 27 | ST831 | 1 | Seafood (Oyster) |
| 19 | 52 | 8 | 110 | 13 | 7 | 22 | 13 | 159 | 14 | ST832 | 1 | Seafood (Oyster) |
| 5 | 78 | *139** | 26 | 17 | 18 | 5 | 8 | 12 | 9 | ST833 | 1 | Seafood (Oyster) |
| *252** | 47 | 14 | 4 | 11 | 23 | 5 | 8 | 5 | 80 | ST834 | 1 | Seafood (Oyster) |
| *252** | 45 | 14 | 6 | 11 | 23 | 5 | 8 | 5 | 80 | ST835 | 1 | Seafood (Oyster) |
| 13 | 25 | 2 | 4 | 13 | 7 | *126** | 18 | *167** | 30 | ST836 | 1 | Seafood (Oyster) |
| 106 | *171** | 64 | *229** | 2 | 60 | 46 | *110** | 17 | 130 | ST837 | 1 | Seafood (Oyster) |
| 2 | 18 | *136** | 98 | 11 | 23 | *123** | 8 | *165** | 18 | ST838 | 1 | Seafood (Oyster) |
| *249** | 25 | 38 | 26 | 73 | 7 | 76 | 48 | *165** | 39 | ST839 | 1 | Seafood (Oyster) |
| 2 | 69 | 2 | 17 | 11 | 23 | 22 | 13 | 5 | 86 | ST840 | 1 | Seafood (Oyster) |
| 5 | *172** | 122 | *230** | *158** | 24 | *126** | 3 | 15 | *173** | ST841 | 1 | Seafood (Oyster) |
| 184 | *167** | 60 | 69 | 11 | *132** | 13 | 89 | *167** | 168* | ST842 | 1 | Seafood (Oyster) |
| 10 | 18 | *136** | 178 | 11 | 23 | *123** | 8 | *165** | 45 | ST843 | 1 | Seafood (Oyster) |
| 19 | 78 | *138** | 65 | 11 | *133** | 95 | 19 | *168** | 18 | ST844 | 1 | Seafood (Oyster) |
| 116 | 8 | 122 | 1 | 26 | 24 | 25 | 3 | 5 | 44 | ST845 | 1 | Seafood (Oyster) |
| 19 | 47 | 14 | 17 | 2 | 7 | 16 | 8 | *166** | 26 | ST846 | 1 | Seafood (Oyster) |
| 10 | 18 | *136** | 98 | 11 | 23 | *123** | 8 | 5 | 45 | ST847 | 1 | Seafood (Oyster) |
| 164 | *168** | 2 | 183 | *152** | *134** | 21 | 89 | 5 | 15 | ST848 | 1 | Seafood (Oyster) |
| 175 | 23 | 14 | 18 | 17 | 17 | 21 | 8 | 4 | 9 | ST849 | 1 | Seafood (Oyster) |
| 34 | 52 | 100 | 69 | 10 | 18 | 76 | 8 | 5 | *169** | ST850 | 1 | Seafood (Oyster) |
| 170 | 23 | 14 | 18 | 17 | 17 | 21 | 8 | 17 | 9 | ST851 | 1 | Clinical (Blood) |
| 111 | 18 | *136** | 98 | 11 | 23 | 76 | 8 | *165** | 45 | ST852 | 1 | Seafood (Oyster) |
| *258** | 8 | 16 | 57 | 2 | 7 | 71 | 8 | 17 | 9 | ST853 | 1 | Clinical (Blood) |
| 111 | 47 | 14 | 62 | 11 | 23 | 5 | 8 | 5 | 80 | ST854 | 1 | Seafood (Oyster) |
| 36 | *177** | 42 | *235** | 11 | 7 | *129** | 16 | *174** | 30 | ST855 | 1 | Seafood (Oyster) |
| *260** | 54 | 1 | 81 | 12 | 4 | 48 | 46 | 66 | 7 | ST856 | 1 | Clinical (Blood) |
| 100 | 17 | 68 | 6 | 73 | 7 | 108 | 95 | 125 | 30 | ST857 | 1 | Seafood (Oyster) |
| 184 | 45 | 62 | 26 | 11 | 22 | 125 | 3 | 53 | 87 | ST858 | 1 | Seafood (Clam) |
| 10 | 18 | *136** | 110 | 11 | 23 | *123** | 53 | *165** | 45 | ST859 | 1 | Seafood (Clam) |
| 34 | 52 | 28 | 60 | 11 | 23 | 21 | 13 | 5 | 52 | ST860 | 1 | Seafood (Oyster) |
| 18 | 27 | 95 | *223** | 20 | *135** | *124** | 8 | 17 | 9 | ST861 | 1 | Seafood (Oyster) |
| 111 | 33 | 8 | 62 | 11 | 2 | 5 | 3 | 53 | 30 | ST862 | 1 | Seafood (Oyster) |
| 5 | 14 | 6 | *224** | 20 | 7 | 18 | 8 | 28 | 9 | ST863 | 1 | Seafood (Oyster) |
| 34 | 125 | 14 | 53 | 2 | 3 | 21 | 59 | *176** | 9 | ST864 | 1 | Seafood (Oyster) |
| 230 | 82 | 102 | 1 | 11 | 7 | 76 | 9 | *166** | 18 | ST865 | 1 | Seafood (Oyster) |
| 107 | 45 | 2 | 112 | 2 | 2 | 20 | 8 | 17 | 43 | ST866 | 1 | Seafood (Oyster) |
| 100 | 8 | *137** | 53 | 2 | *131** | 109 | *109** | 9 | 78 | ST867 | 1 | Seafood (Oyster) |
| 223 | 3 | 18 | 87 | 3 | *137** | 10 | 4 | 43 | 61 | ST868 | 1 | Clinical (Wound) |
| 8 | 5 | *147** | 9 | 8 | 8 | 1 | 1 | 10 | 7 | ST869 | 1 | Clinical (Blood) |
| 34 | 82 | 28 | 60 | 6 | 23 | 109 | 8 | 5 | 52 | ST870 | 1 | Seafood (Oyster) |
| 19 | 47 | 41 | *226** | 2 | *138** | 15 | 8 | 5 | 74 | ST871 | 1 | Seafood (Oyster) |
| 19 | 47 | 14 | *222** | 2 | 7 | 16 | 8 | *166** | 74 | ST872 | 1 | Seafood (Oyster) |
| 2 | 45 | 2 | 62 | 11 | 7 | 8 | 13 | 53 | 86 | ST873 | 1 | Seafood (Oyster) |
| 107 | 135 | 110 | 17 | 26 | 7 | 5 | 8 | 5 | 30 | ST874 | 1 | Seafood (Oyster) |
| *258** | *172** | 38 | *225** | *155** | 23 | *128** | 9 | 14 | 18 | ST875 | 1 | Seafood (Oyster) |
| 230 | 82 | 102 | 110 | 11 | 2 | 5 | 3 | *166** | 18 | ST876 | 1 | Seafood (Oyster) |
| 5 | 79 | 9 | *233** | 21 | 23 | 104 | 16 | 14 | 9 | ST877 | 1 | Clinical (Blood) |
| 34 | 52 | 28 | 75 | 11 | 23 | 21 | 8 | 5 | 88 | ST878 | 1 | Seafood (Oyster) |
| *259** | 33 | 2 | 1 | 11 | 2 | 5 | 3 | 53 | 86 | ST879 | 1 | Seafood (Oyster) |
| 107 | 10 | 2 | 26 | 20 | 2 | 71 | 8 | 17 | *174** | ST880 | 1 | Seafood (Oyster) |
| 22 | 26 | 23 | 65 | *157** | 24 | 13 | 8 | 12 | 125 | ST881 | 1 | Clinical (Blood) |
| *261** | 7 | 25 | 55 | 11 | 7 | *131** | 9 | *175** | 18 | ST882 | 1 | Seafood (Oyster) |
| *252** | 47 | 14 | 110 | 11 | 23 | 5 | 8 | 5 | 52 | ST883 | 1 | Seafood (Oyster) |
| 111 | 52 | 88 | 69 | 10 | 18 | *130** | *112** | 136 | *169** | ST884 | 1 | Clinical (Blood) |
| 191 | 69 | 42 | 1 | 2 | 7 | 23 | 8 | 14 | 125 | ST885 | 1 | Seafood (Oyster) |
| 2 | 82 | 14 | *236** | 11 | 23 | 15 | 13 | 5 | 18 | ST886 | 1 | Seafood (Oyster) |
| 111 | *175** | *144** | 26 | 9 | 7 | *127** | 3 | *171** | 118 | ST887 | 1 | Seafood (Oyster) |
| 111 | 33 | 2 | 62 | 11 | 2 | 5 | 3 | 14 | 30 | ST888 | 1 | Seafood (Oyster) |
| 26 | *173** | 3 | 29 | *153** | 19 | 28 | 20 | 43 | 7 | ST889 | 1 | Clinical (Blood) |
| 230 | 82 | *145** | 110 | 13 | 7 | 76 | 9 | 5 | 18 | ST890 | 1 | Seafood (Oyster) |
| 34 | 25 | *146** | *232** | *156** | 2 | 46 | *111** | 53 | 50 | ST891 | 1 | Seafood (Oyster) |
| 46 | 69 | 2 | *234** | 11 | 7 | 22 | 13 | 12 | 26 | ST892 | 1 | Seafood (Oyster) |
