## Supplemental Table 3 for "Population Structure, Novel Sequence Types, and Antimicrobial Resistance in *Vibrio parahaemolyticus* and *Vibrio vulnificus*: A Whole Genome Sequencing Study of Clinical and Seafood Isolates in New Jersey"

**Supplemental Table 3**: New Jersey Vibrio parahaemolyticus sequences available from the Sequence Read Archive (SRA)

| SRR13968772 | SRR14239729 | SRR18270796 | SRR23285917 | SRR28258081 |
| --- | --- | --- | --- | --- |
| SRR14078458 | SRR14239730 | SRR18326276 | SRR23285928 | SRR28258082 |
| SRR14078460 | SRR14239731 | SRR18326277 | SRR24660882 | SRR28258203 |
| SRR14078461 | SRR14239732 | SRR18326278 | SRR24767365 | SRR28258339 |
| SRR14078462 | SRR14239733 | SRR18326279 | SRR24860854 | SRR28258392 |
| SRR14078463 | SRR14239734 | SRR18326280 | SRR24860855 | SRR28258710 |
| SRR14078464 | SRR14253899 | SRR18326283 | SRR24860856 | SRR28312011 |
| SRR14078465 | SRR14253900 | SRR18326286 | SRR24860863 | SRR28314581 |
| SRR14078466 | SRR14253901 | SRR18326287 | SRR24860864 | SRR28397844 |
| SRR14078523 | SRR14253902 | SRR18326288 | SRR24860890 | SRR28526966 |
| SRR14078616 | SRR14253903 | SRR18326289 | SRR24866247 | SRR29347409 |
| SRR14078728 | SRR14253904 | SRR18326291 | SRR24866293 | SRR29622526 |
| SRR14078783 | SRR14253905 | SRR18326293 | SRR24866469 | SRR29950808 |
| SRR14078891 | SRR14253906 | SRR18326294 | SRR24866473 | SRR29950809 |
| SRR14083589 | SRR15838430 | SRR18347903 | SRR24938687 | SRR29992241 |
| SRR14083599 | SRR17772387 | SRR18490902 | SRR24938688 | SRR30086261 |
| SRR14083601 | SRR17772388 | SRR18709553 | SRR24939838 | SRR30172688 |
| SRR14083603 | SRR17772389 | SRR18709555 | SRR24939894 | SRR30189164 |
| SRR14083604 | SRR17772390 | SRR18709557 | SRR24968887 | SRR30476983 |
| SRR14083605 | SRR17772391 | SRR18709560 | SRR24968888 | SRR30840276 |
| SRR14083606 | SRR17772392 | SRR20106138 | SRR24968889 | SRR30900030 |
| SRR14083608 | SRR17772393 | SRR20334240 | SRR24968890 | SRR32311218 |
| SRR14083609 | SRR17772394 | SRR20334242 | SRR24968891 | SRR32311394 |
| SRR14083610 | SRR17772395 | SRR20630896 | SRR24968894 | SRR32311421 |
| SRR14083612 | SRR17772396 | SRR20751882 | SRR24968895 | SRR32311427 |
| SRR14083613 | SRR17772397 | SRR21091440 | SRR24968896 | SRR32311428 |
| SRR14083614 | SRR17772398 | SRR21091441 | SRR24968897 | SRR32311429 |
| SRR14083615 | SRR17772399 | SRR21202328 | SRR24970290 | SRR32311802 |
| SRR14097072 | SRR17772400 | SRR21443936 | SRR24973787 | SRR32311823 |
| SRR14097073 | SRR17772401 | SRR21582769 | SRR25253060 | SRR32311824 |
| SRR14097075 | SRR17772402 | SRR21658674 | SRR25402846 | SRR32311991 |
| SRR14097077 | SRR17772403 | SRR21863753 | SRR25445515 | SRR32312039 |
| SRR14097080 | SRR17772408 | SRR22055083 | SRR25445516 | SRR32312067 |
| SRR14097081 | SRR17838827 | SRR22055113 | SRR25596844 | SRR32312262 |
| SRR14097082 | SRR17838887 | SRR22055114 | SRR25596851 | SRR32312264 |
| SRR14097085 | SRR17838891 | SRR22055115 | SRR25596853 | SRR32464014 |
| SRR14097086 | SRR17838914 | SRR22055116 | SRR25596863 | SRR32464029 |
| SRR14097087 | SRR17838938 | SRR22055117 | SRR25685586 | SRR32464057 |
| SRR14097089 | SRR17888655 | SRR22055118 | SRR25917051 | SRR32464075 |
| SRR14097090 | SRR17888656 | SRR22055119 | SRR25958957 | SRR32465391 |
| SRR14097093 | SRR17888657 | SRR22055120 | SRR25960485 | SRR32465392 |
| SRR14097094 | SRR17888658 | SRR22055121 | SRR26093579 | SRR32511889 |
| SRR14097095 | SRR17888659 | SRR22055122 | SRR26221638 | SRR32512387 |
| SRR14097097 | SRR17888660 | SRR22055123 | SRR26221724 | SRR32598358 |
| SRR14097098 | SRR17888661 | SRR22055124 | SRR28257117 | SRR32599621 |
| SRR14097099 | SRR17888662 | SRR22055125 | SRR28257119 | SRR32599753 |
| SRR14097100 | SRR17888663 | SRR22055126 | SRR28257123 | SRR32600076 |
| SRR14097102 | SRR17888664 | SRR22055127 | SRR28257128 | SRR32600130 |
| SRR14097103 | SRR17888665 | SRR22055128 | SRR28257129 | SRR32630241 |
| SRR14097104 | SRR17888666 | SRR22055129 | SRR28257130 | SRR32630269 |
| SRR14097106 | SRR17888667 | SRR22084687 | SRR28257289 | SRR32630274 |
| SRR14097107 | SRR17888668 | SRR22246241 | SRR28257291 | SRR32630287 |
| SRR14097108 | SRR17888671 | SRR22246242 | SRR28257506 | SRR32630320 |
| SRR14097109 | SRR18270328 | SRR22333360 | SRR28257781 | SRR32630329 |
| SRR14099159 | SRR18270575 | SRR22356377 | SRR28257846 | SRR32630937 |
| SRR14143451 | SRR18270662 | SRR22356378 | SRR28257849 | SRR32630979 |
| SRR14239725 | SRR18270663 | SRR22704972 | SRR28257912 | SRR32631097 |
| SRR14239726 | SRR18270724 | SRR22704973 | SRR28258075 |  |
| SRR14239727 | SRR18270786 | SRR23285867 | SRR28258078 |  |
| SRR14239728 | SRR18270787 | SRR23285868 | SRR28258080 |  |
