## Supplemental Table 4 for "Population Structure, Novel Sequence Types, and Antimicrobial Resistance in *Vibrio parahaemolyticus* and *Vibrio vulnificus*: A Whole Genome Sequencing Study of Clinical and Seafood Isolates in New Jersey"

**Supplemental Table 4**: New Jersey *Vibrio vulnificus* sequences available from the Sequence Read Archive (SRA)

| SRR13968771 | SRR17838857 | SRR22356373 | SRR28257292 |
| --- | --- | --- | --- |
| SRR14082677 | SRR17838875 | SRR22356375 | SRR28257293 |
| SRR14082678 | SRR17888669 | SRR22356380 | SRR28257509 |
| SRR14082679 | SRR17888670 | SRR22704971 | SRR28257541 |
| SRR14082680 | SRR17888672 | SRR22704974 | SRR28257556 |
| SRR14084253 | SRR17888673 | SRR22704975 | SRR28257750 |
| SRR14084254 | SRR17888674 | SRR23285927 | SRR28257850 |
| SRR14084255 | SRR17888675 | SRR23285934 | SRR28257921 |
| SRR14084256 | SRR17888676 | SRR24860964 | SRR28258043 |
| SRR14084257 | SRR17888677 | SRR24860967 | SRR28258079 |
| SRR14084258 | SRR18270794 | SRR24860970 | SRR28258206 |
| SRR14084259 | SRR18270795 | SRR24860971 | SRR28258360 |
| SRR14084260 | SRR18326229 | SRR24866317 | SRR28305117 |
| SRR14097111 | SRR18326290 | SRR24938627 | SRR28305121 |
| SRR14097113 | SRR18326292 | SRR24938634 | SRR28305157 |
| SRR14097114 | SRR18326302 | SRR24938642 | SRR28305158 |
| SRR14097118 | SRR18326303 | SRR24939220 | SRR28305189 |
| SRR14143403 | SRR18326545 | SRR24939243 | SRR28307656 |
| SRR14143405 | SRR18326546 | SRR24968910 | SRR28307752 |
| SRR14143408 | SRR18326587 | SRR24968917 | SRR28440096 |
| SRR14143457 | SRR18326588 | SRR24968922 | SRR28642447 |
| SRR14143459 | SRR18326589 | SRR24968924 | SRR29622527 |
| SRR14253909 | SRR18344824 | SRR24970394 | SRR30162117 |
| SRR14253910 | SRR18348177 | SRR25004906 | SRR30527401 |
| SRR14253912 | SRR18557333 | SRR25004908 | SRR32311395 |
| SRR14253913 | SRR18709556 | SRR25445517 | SRR32311459 |
| SRR14253914 | SRR18709558 | SRR25495861 | SRR32311836 |
| SRR14253915 | SRR18709559 | SRR25666302 | SRR32311979 |
| SRR14253916 | SRR18730490 | SRR26826440 | SRR32311987 |
| SRR14253923 | SRR18730491 | SRR26942142 | SRR32312065 |
| SRR14253924 | SRR20668340 | SRR28257106 | SRR32312066 |
| SRR14253926 | SRR20751883 | SRR28257109 | SRR32312070 |
| SRR14253927 | SRR21201254 | SRR28257110 | SRR32312263 |
| SRR17772404 | SRR22055156 | SRR28257112 | SRR32464028 |
| SRR17772405 | SRR22055157 | SRR28257113 | SRR32464030 |
| SRR17772406 | SRR22055158 | SRR28257114 | SRR32464074 |
| SRR17772407 | SRR22055159 | SRR28257120 | SRR32465520 |
| SRR17772409 | SRR22055160 | SRR28257121 | SRR32599573 |
| SRR17772410 | SRR22356328 | SRR28257124 |  |
| SRR17772411 | SRR22356329 | SRR28257127 |  |
